# Antenatal care coverage in a low-resource setting: estimations from the Birhan Cohort

**DOI:** 10.1101/2023.04.20.23288874

**Authors:** Clara Pons-Duran, Delayehu Bekele, Sebastien Haneuse, Bezawit Mesfin Hunegnaw, Kassahun Alemu, Munir Kassa, Yifru Berhan, Frederick G.B. Goddard, Lisanu Taddesse, Grace J. Chan

## Abstract

Antenatal care (ANC) coverage estimates commonly rely on self-reported data, which may carry biases. Leveraging prospectively collected longitudinal data, this study aimed to estimate the coverage of ANC, minimizing assumptions and biases due to self-reported information and describing retention patterns in ANC in rural Amhara, Ethiopia. This is a cohort study using data from the Birhan Health and Demographic Surveillance System and its nested pregnancy and birth cohort, the Birhan Cohort. The study population were women enrolled and followed during pregnancy between December 2018 and April 2020. ANC visits were measured by prospective facility chart abstraction and self-report at enrollment. The primary study outcomes were the total number of ANC visits attended during pregnancy and the coverage of at least one, four and eight ANC visits. Additionally, we estimated ANC retention patterns.We included 2069 women, of which 150 (7.2%) women enrolled <13 weeks of gestation with complete prospective facility reporting. Among 150 women, ANC coverage of at least one visit was 97.3%, whereas coverage of four visits or more was 34.0%. Among all women, coverage of one ANC visit was 92.3%, while coverage of four or more visits was 28.8%. No women were found to have attended eight or more ANC visits. On retention in care, 70.3% of participants who had an ANC visit between weeks 28 and <36 of gestation did not return for a subsequent visit. Despite the high proportion of pregnant women who accessed ANC at least once in our study area, the coverage of four visits remains low. Further efforts are needed to enhance access to more ANC visits, retain women in care, and adhere to the most recent National ANC guideline. It is essential to identify the factors that lead a large proportion of women to discontinue ANC follow-up.

## Introduction

Antenatal care (ANC) coverage is an indicator of utilization of essential health services for pregnant women, defined as the proportion of women with a birth in a certain period who attended a specific number of ANC visits for their most recent pregnancy [1, 2]. ANC provides an opportunity to anticipate complications, deliver preventive strategies, and ultimately identify population groups at high risk of experiencing adverse events during pregnancy to prevent them [1]. ANC could contribute to reducing maternal and neonatal mortality figures and improve other maternal and newborn health outcomes [3, 4]. Ethiopia is the second most populous country in Africa, with the fastest growing economy on the continent [5]. Despite an increasing trend in ANC coverage over the past two decades [6], the coverage of at least one ANC visit during pregnancy was lower than 75%, and the proportion of women who remain in care for up to four or more visits was 43% in 2019 [7]. These country estimates are far from the national target of achieving 81% coverage of four or more ANC visits by 2025 [8]. Recently in early 2022, the National ANC guideline was updated, and now a minimum of eight ANC visits during pregnancy are recommended [9].

There is evidence suggesting that self-reports of ANC use are largely biased when coverage is low and those self-reports have low specificity [10]. This may be due to long recall periods, social desirability bias, or poor understanding of what constitutes ANC. In Ethiopia and other low-resource settings, there is a lack of studies that validate self-reported data about ANC use, and the reliability of those estimates is unknown. Further, most studies on ANC coverage conducted in Ethiopia are cross-sectional [11-16], and to our knowledge, there is no evidence of ANC estimates based on longitudinal prospective data or direct observation. In addition to the potential biases that self-reported data may imply, cross-sectional studies make it difficult to investigate retention in care patterns and timing of the recommended ANC visits.

Quality and accurate estimates of ANC coverage from different regions in Ethiopia are fundamental to understanding the level of ANC use at local level and to tracking implementation progress of the recently launched National ANC guideline [9]. Reliable estimates are needed to inform the design and delivery of interventions to enhance ANC attendance. This study presents an opportunity to leverage prospectively collected longitudinal data from a maternal and child cohort. We aimed to estimate the coverage of ANC using an approach that minimizes assumptions and biases from self-reported information and to describe retention patterns in ANC in rural Amhara, Ethiopia.

## Methods

### Study design and setting

We conducted a cohort study in the Birhan field site, including 16 villages in Amhara Region, Ethiopia, covering a mid-year population of 77,766, to estimate morbidity and mortality outcomes among 17,108 women of reproductive age and 8,554 children under-five with house-to-house surveillance every three months. The field site includes a health and demographic surveillance system (HDSS), the Birhan HDSS, a platform for community and facility-based research and training that was established in 2018, with a focus on maternal and child health [17]. Nested in the site is an open pregnancy and birth cohort, the Birhan Cohort, that enrolls approximately 2,000 pregnant women and their newborns per year with rigorous longitudinal follow-up over the first two years of life and household data linked with health facility information [18]. The catchment area is rural and semi-urban, covers both highland and lowland areas, and includes two different districts, Angolela Tera, and Kewet/Shewa Robit.

We used data from the Birhan HDSS and Birhan Cohort to estimate coverage of ANC-related indicators [17, 18]. The HDSS provides estimates and trends of health and demographic outcomes including morbidity among women of reproductive age and children under two years and births, deaths, marriages, and migration in the entire population [17]. The pregnancy and birth cohort, generates evidence on pregnancy, birth, and child outcomes using clinical and epidemiological data at both the community and health facility level [18].

### Study population

We used data of women enrolled and followed until delivery in the Birhan Cohort, both at the facility and community level, between December 2018 and April 2020. We excluded participants with abortions or miscarriages and implausible documented gestational ages at enrollment (≤0 weeks or ≥46 weeks) and/or delivery (<28 weeks or ≥46 weeks). Gestational age was estimated using the best available method from ultrasound measurements, date of last menstrual period, fundal height, and maternal recall of gestational age in months [19].

### Study outcomes

ANC visits were measured by prospective facility chart abstraction from enrollment and self-report at enrollment. In a subgroup of women, retrospective facility chart abstraction was done. We defined ANC coverage as the proportion of women enrolled in the Birhan Cohort who attended ANC visits during pregnancy. The primary study outcomes were the total number of ANC visits attended during pregnancy and the coverage of at least one, four, and eight ANC visits.

Secondary study outcomes included retention in care, defined as the continued engagement in facility ANC. We defined those secondary outcomes as the proportion of study participants in care at different gestational age times (ANC1 window, <16 weeks; ANC2 window, 16 - <28 weeks; ANC3 window, 28 - <36 weeks; and ANC4 window, ≥36 weeks) and the proportion of participants in care who were lost after each of those time windows.

### Analysis

We estimated ANC coverage by adding the number of prospectively recorded visits from enrollment to the number of self-reported visits at enrollment. Since study participants were enrolled at different times during pregnancy, we estimated ANC coverage among the cohort of women who enrolled <13 weeks where most of the ANC visits were recorded prospectively to minimize recall bias. In addition, we also estimated ANC coverage for the entire population. For coverage outcomes, descriptive statistics were undertaken; frequencies, proportions, and Agresti-Coull 95% confidence intervals (CI)[20] were reported for dichotomous outcomes, while median and interquartile range (IQR) were used to report continuous outcomes.

We assessed the quality and reliability of self-reports by comparing the counts of self-reported visits at enrollment and the retrospectively collected visits from charts for a subset of participants. More details of this analysis can be found in the S1 File.

To assess comparability between the study subsample (women enrolled <13 weeks) and the remaining cohort participants, we compared the socio-demographic characteristics and obstetric history of participants enrolled <13 weeks and ≥13 weeks of gestation using descriptive statistics and chi-squared tests, t-test, and Fisher exact tests. Further, women enrolled in the cohort at different gestation age weeks were compared in terms of access to ANC services during second and third trimesters using frequencies and proportions of attendance to at least one ANC visit at different time windows.

To investigate secondary study outcomes, the proportion of participants in care during the window times for the different ANC visits was represented in an alluvial plot. Proportions of women in care during ANC1 to ANC4 window times were estimated for the cohort of women enrolled <13 weeks. To calculate retention, the proportion of participants lost after each ANC visit was estimated using the total number of women in care for that visit as a denominator. The proportion of women lost after ANC3 did not include those who delivered before gestational week 36. Descriptive statistics were undertaken, with frequencies, proportions, and Agresti-Coull 95% CI reported [20].

To investigate the potential selection bias due to loss to follow-up, we conducted a sensitivity analysis estimating coverage of ANC in the missing data under several scenarios each of which assume that ANC coverage among lost women would be lower than that of women followed until delivery. Specifically, for the outcome ‘at least one ANC visit’, we assumed that all women who did not have a visit before being lost never had any ANC visits. For the outcome ‘four ANC visits or more’, we considered three scenarios where women lost to follow-up had ANC coverage in a range from 0% to 90% of the ANC coverage estimated among women who were not lost to follow-up based on the number of visits attended when they were lost. More details of these scenarios can be found in the S2 File.

Analysis was conducted using R version 4.2.2, and Stata version 17.

#### Ethical considerations

Ethical clearance was obtained from the Ethics Review Board (IRB) of Saint Paul’s Hospital Millennium Medical college, (Addis Ababa, Ethiopia) [PM23/274], Boston Children’s Hospital (Boston, United States) [IRB-P00028224], and Harvard T.H. Chan School of Public Health (Boston, United states) [IRB19-0991]. Signed informed consent was obtained from all participants.

Authors had no access to participants’ identifiable information during or after data collection unless they were involved in field data collection activities and data quality assurance. All study procedures were followed per protocol and participants confidentiality and anonymity was ensured.

## Results

A total of 2303 women were enrolled in the MCH cohort and delivered during the time frame of the study. We excluded 95 (4.1%) women due to pregnancy losses (62, 2.7%) and implausible gestational ages (33, 1.4%). Among the remaining 2208 women, 139 (6.3%) were lost to follow-up; 2069 women were included in the analysis, of which 150 (7.2%) were enrolled in the cohort before 13 weeks of gestation. The participant characteristics for the groups of women enrolled <13 weeks and ≥13 weeks of gestation were similar (Table 1). There were no statistically significant differences observed for age, residency, socio-economics, obstetric history, and timing of ANC visits (see S3 File).

**Table 1.** Characteristics of the study population and by gestational age at enrollment.

| Variables | Enrolled <13 weeks |  | Enrolled ≥13 weeks |  |  | Total |  |
| --- | --- | --- | --- | --- | --- | --- | --- |
|  | (N=150) |  | (N=1919) |  |  | (N=2069) |  |
|  | N | mean (SD) | N | mean (SD) | p-value | N | mean (SD) |
| Age at conception (years) | 150 | 27.9 (6.4) | 1919 | 27.0 (6.1) | 0.12 | 2069 | 27.1 (6.1) |
| Walking time to nearest health facility (minutes) | 150 | 66.2 (57.6) | 1909 | 68.1 (54.7) | 0.69 | 2059 | 68.0 (54.9) |
|  | **N** | **n (%)** | **N** | **n (%)** | **p-value** | **N** | **n (%)** |
| Woreda | 150 |  | 1915 |  | 0.44 | 2065 |  |
| Angolela Tera |  | 62 (41.3) |  | 861 (45.0) |  |  | 923 (44.7) |
| Kewet |  | 88 (58.7) |  | 1054 (55.0) |  |  | 1142 (55.3) |
| Cannot read and write | 150 | 72 (48.0) | 1914 | 866 (45.2) | 0.57 | 2064 | 938 (45.4) |
| Wealth index | 150 |  | 1919 |  | 0.38 | 2069 |  |
| 1<sup>st</sup> quintile (the wealthiest) |  | 36 (24.0) |  | 391 (20.4) |  |  | 427 (20.6) |
| 2<sup>nd</sup> quintile |  | 28 (18.7) |  | 447 (23.3) |  |  | 475 (23.0) |
| 3<sup>rd</sup> quintile |  | 33 (22.0) |  | 386 (20.1) |  |  | 419 (20.3) |
| 4<sup>th</sup> quintile |  | 30 (20.0) |  | 328 (17.1) |  |  | 358 (17.3) |
| 5<sup>th</sup> quintile (the least wealthy) |  | 23 (15.3) |  | 367 (19.1) |  |  | 390 (18.8) |
| Ethnicity | 150 |  | 1915 |  | 1 | 2065 |  |
| Amhara |  | 138 (92.0) |  | 1748 (91.3) |  |  | 1886 (91.3) |
| Oromo |  | 8 (5.3) |  | 113 (5.9) |  |  | 121 (5.9) |
| Other |  | 4 (2.7) |  | 54 (2.8) |  |  | 58 (2.8) |
| Primiparous | 150 | 42 (28.0) | 1919 | 605 (31.5) | 0.42 | 2069 | 647 (31.3) |
| History of stillbirth | 150 | 13 (8.7) | 1919 | 97 (5.1) | 0.09 | 2069 | 110 (5.3) |
| History of preterm birth | 150 | 6 (4.0) | 1919 | 36 (1.9) | 0.12 | 2069 | 42 (2.0) |

Note: SD – standard deviation

Among the 150 women who enrolled in the cohort before 13 weeks of gestation, 97.3% (95%CI 93.1% - 99.2%) had at least one ANC visit, whereas only 34.0% (95%CI 26.9% - 41.9%) of the women had four or more ANC visits (Table 2). No women attended eight or more ANC visits. Almost a third of women (44, 29.3%) attended exactly three visits (Fig 1).

**Table 2.** ANC coverage outcomes of women enrolled <13 weeks of gestation.

| ANC outcomes | N | Median | IQR |
| --- | --- | --- | --- |
| Number of ANC visits | 150 | 3 | 2-4 |
|  | N | n | % (95% CI) |
| At least one ANC visit | 150 | 146 | 97.3 (93.1 – 99.2) |
| Four or more ANC visits | 150 | 51 | 34.0 (26.9 – 41.9) |
| Eight or more ANC visits | 150 | 0 | 0 |

Note: ANC – antenatal care; CI – confidence interval; IQR – interquartile range

**Fig 1.**
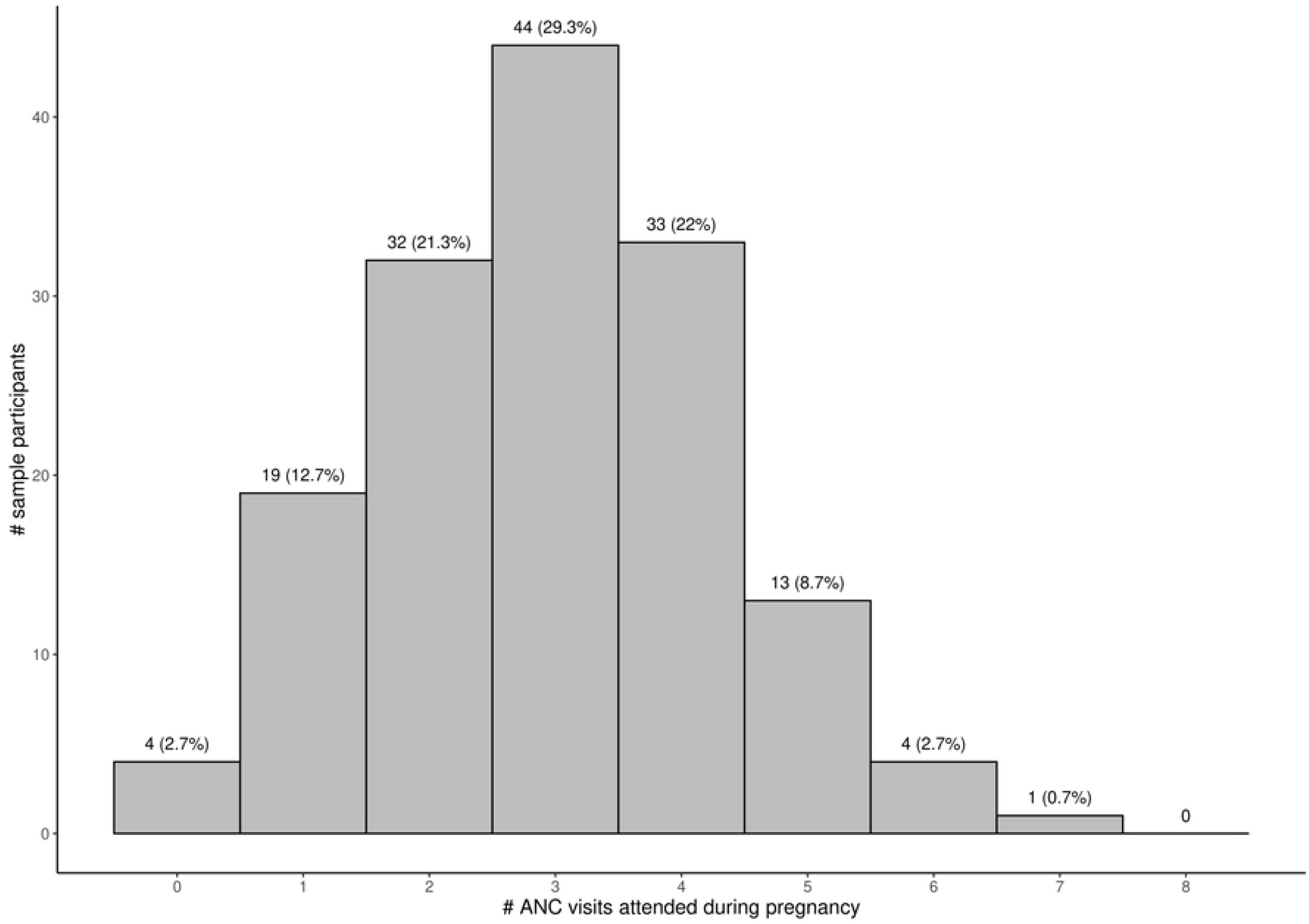
Distribution of ANC visits attended during pregnancy by women enrolled <13 weeks of gestation. Footnote: ANC – antenatal care

In the sensitivity analysis, which estimated ANC coverage in the missing data under several scenarios, there was minimal change in the primary outcomes. Details can be found in the S2 File. Attendance of at least one ANC visit was 96.4% (95%CI 92.2 - 98.5) when assuming all women who did not have a visit before being lost to follow-up never accessed ANC. Coverage of four or more ANC visits ranged between 31.4% (95% CI 24.9 – 38.8%) and 32.9% (95%CI 26.2% - 40.3%) when assuming different ranges of ANC coverage among women who were lost to follow-up.

Among the total population of 2069 women who enrolled at any time during pregnancy, coverage of at least one and four visits was lower (92.3% and 28.8% respectively), and the proportion of individuals who did not attend any ANC visit during pregnancy was higher (7.7%) than for the subsample of women enrolled <13 weeks (S4 File). There was an agreement close to 50% between the number of self-reported visits at enrollment by study participants and the number of visits recorded in charts, suggesting recall bias among self-reported visits, which were predominantly the source of data for retrospective visits for the women who enrolled later in pregnancy (S1 File).

Retention in care, or continued engagement in ANC visits, was low. More than 80% of women were in ANC before week 16 of gestation, and that proportion decreased over time to 29% for the time window of ≥36 weeks (Fig 2). The largest drop-out from ANC occurred after the ANC3 time window (28 to <36 weeks) when 70.3% of study participants who had a visit in that period did not return for a subsequent visit (Table 3).

**Fig 2.**
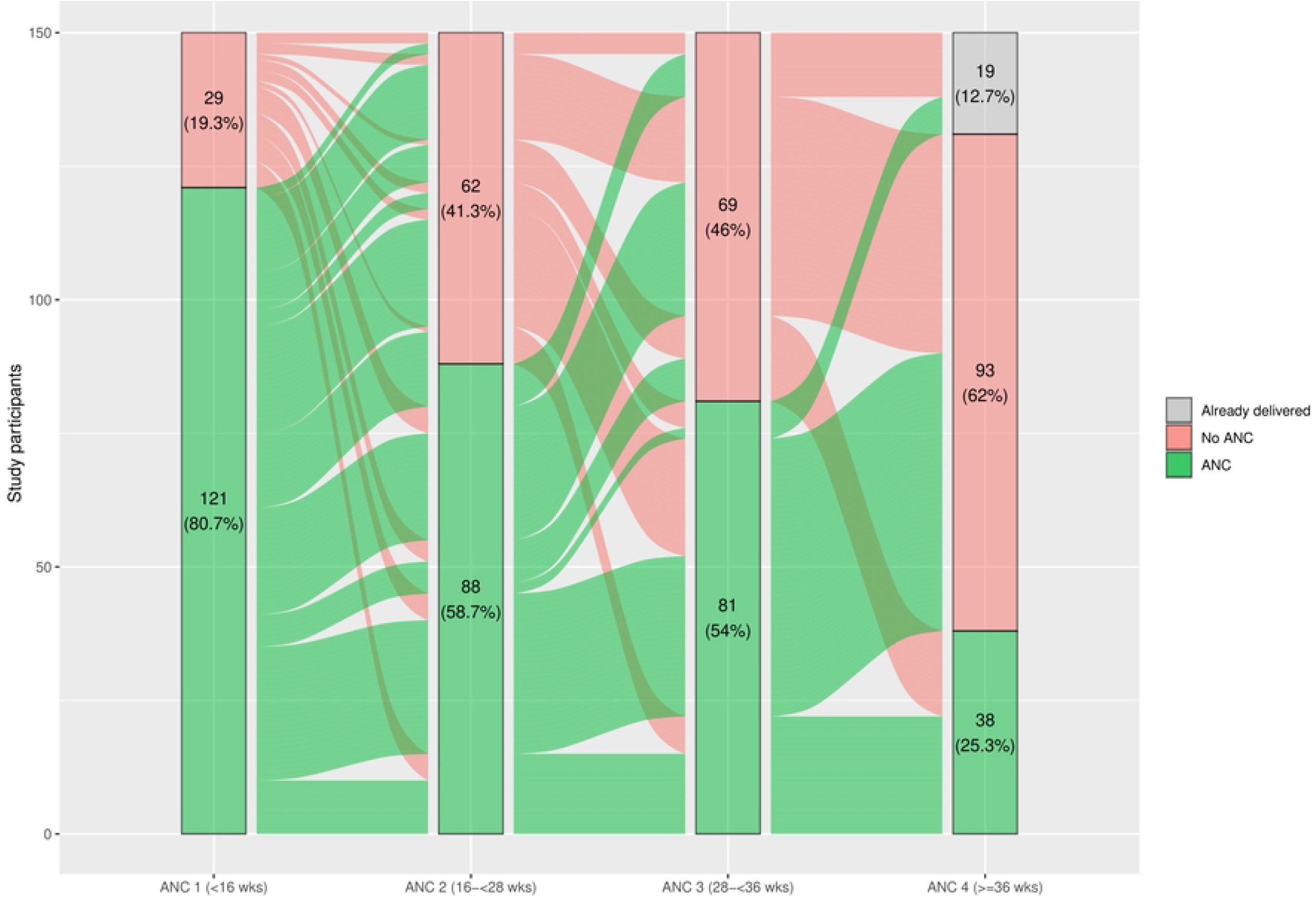
Distribution of ANC visits by window time among women enrolled <13 weeks of gestation. Footnote: ANC – antenatal care

**Table 3.** Retention outcomes of women enrolled <13 weeks of gestation.

| Retention outcomes | N | n | % (95% CI) |
| --- | --- | --- | --- |
| Lost after ANC 1 (<16 weeks) | 121 | 16 | 13.2 (8.2 – 20.5) |
| Lost after ANC 2 (16 to <28 weeks) | 88 | 33 | 37.5 (28.1 – 48.0) |
| Lost after ANC 3 (28 to <36 weeks)* | 74 | 52 | 70.3 (59.0 – 79.5) |

\* The denominator (N) excludes women who delivered before week 36 of gestation
Note: ANC – antenatal care; CI – confidence interval

## Discussion

In this study we found that the coverage of at least one ANC visit during pregnancy is high and close to universal coverage, while coverage of four visits or more remains lower, close to one-third of the sample of women enrolled in the first trimester of pregnancy. No women in our study sample attended eight or more ANC visits. Our findings add valuable information and strengthen the available evidence on ANC coverage that in Ethiopia and other low-resource countries predominantly comes from cross-sectional studies. Longitudinal studies offer more complete sources of information including prospective repeated events, without depending on retrospective self-reported data. We only found cross-sectional studies available in northern Ethiopia to compare with our results. In the Birhan field site between 2018 and 2020, ANC coverage of at least one visit was higher than the results of the 2019 Demographic and Health Survey (DHS) in Amhara region [7, 13]. The Ethiopian mini DHS 2019 reports ANC coverage of pregnancies of the five past years prior to the survey [7]. Thus, it is possible that our coverage of at least one visit is greater due to more attention, policies, and health system resources dedicated to ANC in recent years. On the contrary, our estimates for a minimum of four ANC visits were lower than other studies in Amhara region [7, 13, 15], likely because of the rural nature of the Birhan field site and the potential recall and social desirability bias inherent to the cross-sectional estimates from available studies. No studies reported the coverage of eight or more ANC visits during pregnancy in Ethiopia. Since a minimum of eight ANC visits was not recommended until early 2022, poor coverage of this indicator was expected.

The most common indicator of timely use of ANC services is early ANC attendance, since ANC initiation in the first weeks of pregnancy allows health providers to screen women and conduct tests that are more effective to prevent future complications and assess risks in early weeks [21]. Studies conducted in Ethiopia are not an exception and usually report early ANC attendance as the only indicator of ANC timing [7, 13-15]. However, in order to achieve the recommended number of ANC visits, and to update the information collected in the first visit, timing of subsequent contacts is paramount. To our knowledge, there are no studies on the timing of ANC visits after initiation and on retention in care during pregnancy. Our study demonstrated a trend towards a progressive decrease of ANC attendance during pregnancy, with an especially low proportion of ANC visits after 36 weeks of gestation. The largest drop-out occurred during the window time of the third ANC visit (28 to <36 weeks). The reduced attendance over time may be due to known barriers to ANC utilization that could have a higher impact during the last weeks of pregnancy: negative previous experiences at health facilities, increased fatigue, pregnancy-related signs and symptoms, cultural norms, remoteness that hinders mobility, and lack of family support [6, 22, 23]. In addition, since some ANC components are mainly covered during earlier visits (e.g. complete medical and gynecologic history, ultrasound scan, HIV, hepatitis B and syphilis tests) [9], pregnant women may believe that late visits are less important.

High coverage and continued engagement in ANC are fundamental to reduce preventable pregnancy complications and improve perinatal mortality [3, 4, 24]. In addition, ANC provides the opportunity to be in frequent contact with health providers and improve trust in the health system [25]. ANC attendance is associated with higher rates of facility and skilled-attended delivery [26, 27], which in turn has been observed to decrease perinatal and neonatal mortality compared with home births [28].

Most available ANC coverage estimates from low-resource settings are generated with cross-sectional data and fully based on self-reported information. Our assessment of quality and reliability of self-reported data on ANC visits highlighted a disagreement between data reported by mothers and recorded data in health facility charts, suggesting that self-reports are biased and may decrease the accuracy of estimates. These results are similar to a validation cross-sectional study conducted in rural China that showed large biases in self-reported data of ANC attendance before 12 weeks of gestation by using the ratio of self-reported over recorded coverage [10]. Our study minimized the potential inaccuracy caused by self-reported information and its inherent biases by leveraging a prospective longitudinal cohort study and restricting the study sample to individuals enrolled early in pregnancy. Our findings flag the possibility that many sources of ANC coverage carry biases and errors. In this context, it is necessary to further explore the use of prospectively collected data and utilize existing pregnancy cohorts to estimate ANC-related outcomes. It is necessary to advocate for increased resource allocation for open cohorts and longitudinal studies, and when those data sources are not available, new methodological approaches and mitigation strategies are critical to interpreting and using cross-section data for decision making [29].

Study results need to be interpreted in light of some limitations. Attrition, which is inherent to cohort studies, may have introduced selection bias to the results. However, in a sensitivity analysis, study findings remained consistent and similar across three different scenarios with lost to follow-up data. The small sample size of women who enrolled in the cohort before 13 weeks of gestation to estimate ANC coverage and retention indicators led to wide confidence intervals surrounding our estimates. This trade-off was made to minimize biases caused by self-reported information, leading to more accurate estimates. It was challenging to retrieve data from health facilities outside of the study catchment area, although we suspect the number of women seeking care at facilities outside the catchment were few. While we did not observe any differences in baseline characteristics or timing of ANC between women who enrolled <13 weeks of gestation and those enrolled later in pregnancy, it is possible that there exist unmeasured characteristics between the cohort of women who enrolled <13 weeks compared to the entire cohort sample.

Despite excellent coverage of one or more ANC visits, adequate coverage of the previously recommended four or more ANC visits has not been reached yet in Ethiopia. The recent launch in 2022 of the National ANC guideline to achieve eight or more ANC visits creates an opportunity to continue devoting efforts and resources to increasing adherence and retention of pregnant women in ANC all over the country [9]. As health system and health professionals target adherence to the new ANC guideline, the number of visits attended by pregnant women may potentially experience an increase. As National ANC guidelines aligned with the World Health Organization (WHO) recommendations are being rolled out in Ethiopia and in other low-resource countries, longitudinal studies play an important role in the evaluation and monitoring of implementation through accurate and reliable metrics.

## Conclusion

This study presents a rigorous methodological approach to accurately estimate ANC coverage, leveraging longitudinal data from a pregnancy and birth cohort in Amhara region, Ethiopia. Despite the high proportion of pregnant women who accessed ANC at least once in the study area, further efforts are needed to enhance access to more ANC visits and retain women in care during pregnancy to achieve the global WHO target of eight ANC visits [1]. Furthermore, it is essential to identify and mitigate the barriers that contribute to the large proportion of women who discontinue ANC follow-up after initial ANC visits. Longitudinal prospective data collection should be promoted to minimize the general reliance on self-reports of ANC attendance that could lead to biased estimates and to obtain useful metrics of visits timing to describe patterns of retention in care.

## Data Availability

Data used to produce the results of this article will be publicly available upon publication/acceptance in the following repository: https://dataverse.harvard.edu/dataverse/harvard (we are working on it, we will inform the editor as soon as the data is available online)

## Acknowledgements

We thank all the mothers and children who participated in the study (Birhan HDSS and Birhan Cohort) and the community of the Birhan field site. We also thank data collectors, supervisors, coordinators, and the HaSET team for their contributions.

## Supporting information

**S1 File. Quality and reliability assessment of self-reports**. It includes:

- S1 Table. Distribution of recorded visits that occurred before enrollment and self-reported visits at enrollment for individuals with at least one recorded visit before enrollment

**S2 File. Sensitivity analysis**. It includes:

- S2 Table. Sensitivity analysis: scenarios of ANC coverage of four or more visits

**S3 File. Comparability assessment**. It includes:

- S3 Table. Timing of ANC visits across different gestational ages at enrollment

**S4 File. ANC coverage in the entire Birhan Cohort**. It includes:

- S1 Fig. Distribution of ANC visits attended during pregnancy by women enrolled in the Birhan Cohort
- S4 Table. ANC coverage outcomes of women enrolled in the Birhan Cohort

